# Regional Homogeneity in Muscle Resting-State fMRI as a Novel Marker of Muscle Activation

**DOI:** 10.1101/2025.09.11.25335521

**Authors:** Tereza Hubená, Radim Krupička, Robert Jech

## Abstract

Assessing muscle activity is essential for diagnosis and treatment of neuromuscular disorders such as spasticity and dystonia. While muscle functional magnetic resonance imaging (MRI) enables non-invasive imaging of muscle activation, conventional methods rely on comparisons between rest and activity, which are unsuitable for patients with sustained muscle contractions. This pilot study introduces a novel resting-state fMRI (rs-fMRI) approach using regional homogeneity (ReHo) to evaluate muscle activity without requiring activation paradigms. Eight healthy male participants performed separate isometric plantar and dorsal foot flexion tasks during 3T MRI scanning. rs-fMRI data were analyzed using ReHo to assess local synchronization of BOLD signal. Calf muscle activation was quantified using z-transformed ReHo values and activation thresholds were derived via ROC analysis. Statistical differences in activation between tasks were assessed using the Wilcoxon signed-rank test. Significant ReHo differences were observed between active and inactive muscles during both tasks. This study demonstrates the potential of rs-fMRI combined with ReHo analysis as a non-invasive method for detecting muscle activity using one series of volumes. Further research in larger cohorts is warranted to validate and expand this approach.

## Introduction

Analysis of muscle activity and physiological processes is essential for the diagnosis and treatment of neurological disorders affecting muscles, such as dystonia, spasticity or muscle hyperactivity disorders (Bu et al., 2019; Matsuda et al., 2012; Ricotti et al., 2016; West et al., 2015), as well as for understanding muscle function in rehabilitation, aging, and metabolic conditions (Messer et al., 2020; O’Connor et al., 2006; Dooley et al., 2020; Stacy et al., 2016; Zaeske et al., 2022; Towse et al., 2011).

Muscle functional magnetic resonance imaging (m-fMRI) effectively visualizes changes in oxygenation and blood flow, offering noninvasive insights into muscle activity. It can complement EMG in assessing muscle behaviour and provide access to deeper muscle structures. The key advantage of m-fMRI is its ability to capture an entire limb in a single scan, allowing simultaneous measurement of both deep and superficial muscles (Segal, 2007; Castagna & Albanese, 2019). However, their current application is primarily limited to evaluating the spatial distribution of muscle activation, in contrast to the dynamic assessments provided by EMG techniques (Muller et al., 2016). m-fMRI can quantify perfusion-related signal changes in skeletal muscle during contractions (Wigmore et al., 2004). This technique detects dynamic signal changes caused by local fluctuations in the ratio of oxyhemoglobin to deoxyhemoglobin, known as blood oxygen level-dependent (BOLD) signals (Muller et al., 2016; Stacy et al., 2016; Yamaguchi et al., 2021). A greater negative BOLD response suggests lower deoxyhemoglobin concentration (Muller et al., 2016).

BOLD image quality improves with MRI field strength, with 7T offering better contrast than 3T (Towse et al., 2016), although most studies still use 3T (Bourne et al., 2018; Bourne et al., 2017; Messer et al., 2020) or even 1.5T (Akima et al., 2000; Mendiguchia et al., 2013; O’Connor et al., 2006). Research has primarily targeted the lower limbs—especially calf muscles (Akima et al., 2000; Meyer et al., 2004; Muller et al., 2016a; Towse et al., 2011; Sanchez et al., 2010; Partovi et al., 2012), while some studies analyzed the thigh muscles (Bourne et al., 2018; Messer et al., 2020; West et al., 2015; Bourne et al., 2017; Li et al., 2014; Mendiguchia et al., 2013), foot (O’Connor et al., 2006; Stacy et al., 2016), forearm (Ricotti et al., 2016) and back muscles (Huang et al., 2020).

BOLD is typically assessed using activation maps by subtracting resting from active MRI images, with differences shown directly (O’Connor et al., 2006; Norrbrand et al., 2011) or as percentages relative to resting values (Bourne et al., 2017; Bourne et al., 2018; Messer et al., 2020). However, this approach is not suitable for hyperfunctioning dystonic muscles, where constant contractions prevent resting-active comparisons. In such cases, resting-state fMRI (rs-fMRI) is a viable alternative, enabling analysis without activation tasks. Parameters like regional homogeneity (ReHo) used in brain analysis can assess spontaneous BOLD fluctuations (Zang et al., 2004). ReHo quantifies the similarity or synchronization of the BOLD signal time series among neighboring voxels, reflecting local functional coherence. In the context of skeletal muscle, we hypothesize that during activation, spatially clustered motor units exhibit synchronized hemodynamic responses, which can be captured by increased ReHo values. Thus, ReHo may serve as an indirect marker of muscle activity.

In this study, we applied ReHo to m-fMRI to investigate its potential for detecting muscle activity during isometric contractions, offering a novel noninvasive assessment method. Using rs-fMRI, we captured BOLD signal variations in the calf muscles during plantar and dorsal flexion, eliminating the need for traditional activation paradigms. By applying ReHo, we assessed localized synchronization of BOLD fluctuations, providing a method to evaluate muscle activity without relying on direct comparisons between resting and active images.

## Methods

This pilot study included 8 healthy male probands (mean age 26.7 ± (SD) 6.7 years) with no history of lower limb injury and no motor or neurological disorder. The probands were examined using magnetic resonance imaging (MRI). They were instructed to perform plantar flexion followed by dorsal flexion after a 5-minute relaxation period. The leg position was stabilized from the sides around the ankle and knee to prevent lateral movement, and flexion was voluntary but supported by sandbags around the feet. To ensure sustainability throughout the measurement duration, the flexion was performed at approximately 60% of maximal force rather than full intensity. Muscle group activation was practiced under the supervision of physiotherapists to ensure the correct leg position and posture during the tasks.

MRI data covering a 10 cm section of the main calf region, including the area with the largest muscle volume, were acquired using 3T magnetic resonance system MAGNETOM Skyra (Siemens). For rs-fMRI, BOLD EP2D volumes were acquired (TR: 2000 ms, TE: 30 ms, 304 volumes, 64×64×30 voxels, voxel size: 3×3×3.45 mm, FA: 90 deg). Additionally, proton density weighted (PD) TSE volumes (TR: 6380 ms, TE: 41 ms, 320×260×60 voxels, voxel size 0.6×0.6×2.5 mm, FA: 150 deg) were acquired for anatomical reference. Anatomical references were acquired separately for both plantar and dorsal flexion conditions.

Anatomical volumes were used to manually segment five muscle compartments using ITK-SNAP software (Yushkevich et al., 2006). Two of the selected muscles are primarily active during dorsal flexion (tibialis anterior - TA, extensor digitorum longus - EDL), and three are involved in plantar flexion (peroneus longus - PER, soleus - SO, lateral and medial gastrocnemius - MG, LG). Selected muscles and segmentation in one slice of the calf volume are shown in Figure 1.

**Figure 1.**
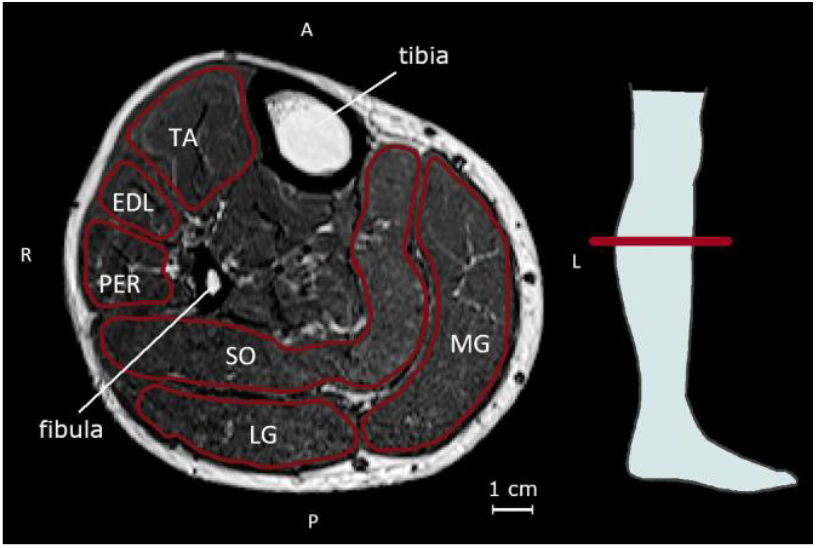
Segmentation of calf muscle. The red line marks the segmented muscle regions used for functional MRI analysis. The red line on the drawing of the leg indicates the position of the displayed cross-section. (TA – tibialis anterior, EDL – extensor digitorum longus, PER – peroneus longus, SO – soleus, LG – lateral gastrocnemius, MG – medial gastrocnemius; A-anterior, P-posterior, L-left, R-right)

Functional volumes were registered with anatomical volumes across the entire calf region. The first 10 seconds of each time series were discarded to ensure the signal reached a steady state. ReHo was calculated for each voxel using a cluster size of 27 voxels, then smoothed with a 3×3×3 voxel mean filter. The resulting ReHo values were transformed into z-score ReHo (zReHo) by normalizing them - subtracting the mean and dividing by the standard deviation within the masked ReHo region. Muscle activation was quantified by calculating the percentage of activated voxels for each segmented muscle. A voxel was considered activated if its zReHo value exceeded the predefined threshold, which was computed using Receiver Operating Characteristic (ROC) analysis. For each participant, an individual ROC curve was generated by comparing zReHo values in muscles expected to be active during the given task (TA and EDL for dorsal flexion; PER, SO, MG, and LG for plantar flexion) against those in other segmented muscles. These curves evaluated the sensitivity and specificity of different zReHo thresholds in distinguishing active from inactive regions. The individual ROC curves were then averaged to derive a single aggregated ROC curve across all subjects, from which the optimal activation threshold was identified. This processing pipeline is shown in Figure 2.

**Figure 2.**
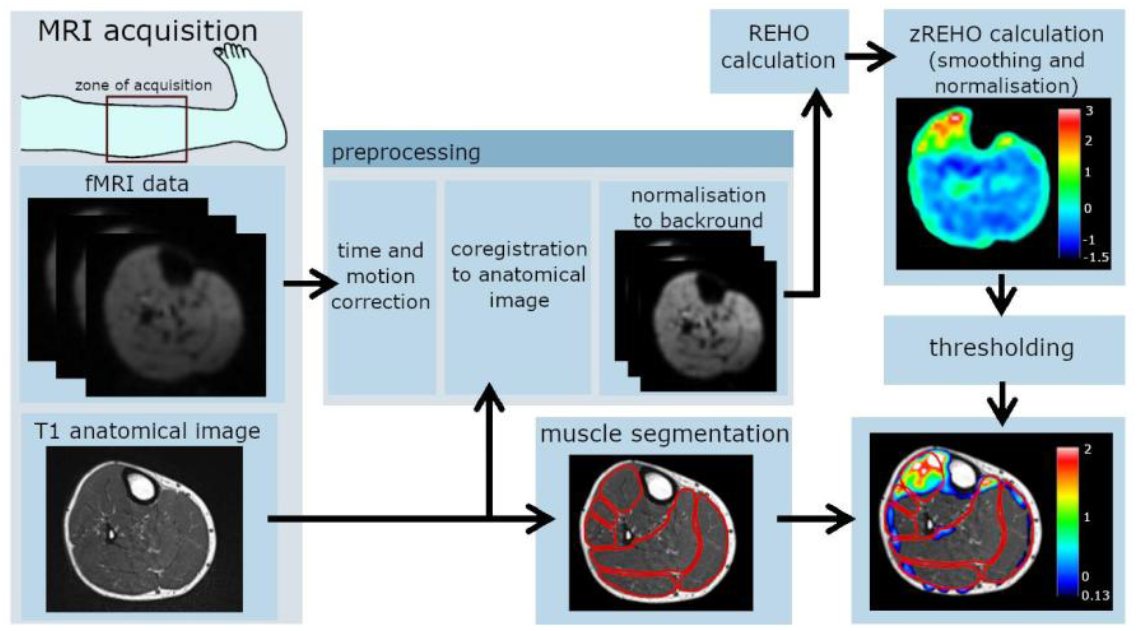
Diagrammatic illustration of the MRI processing pipeline on a representative slice; applied to the full volume. zReHo is calculated from preprocessed functional volumes and displayed over segmented anatomical volumes. The threshold of displayed zReHo values was set to 0.13 based on ROC analysis.

Activated voxels were identified based on the defined threshold. To compare the percentage of activated voxels in each segmented muscle during dorsal flexion versus plantar flexion, a pairwise Wilcoxon signed-rank test was used for statistical analysis. The significance level was set at 0.05 and corrected to 0.008 using the Bonferroni correction.

Results were also visually inspected across subjects and separately for each task. For the analysis, MATLAB (The Mathworks, Natick, Massachusetts, USA) with SPM12 (Statistical Parametric Mapping, Friston et al., 2007) and DPARSF (Data Processing Assistant for Resting-State fMRI, Yan et al., 2016) toolboxes were used.

The study was approved by the Ethics Committee of the General University Hospital in Prague, all participants were properly informed and signed the informed consent form.

## Results

The regional homogeneity results are displayed on an anatomical volume with segmented muscle groups for each subject. The ROC curve of zReho values with the calculated threshold are shown in Figure 3. The optimal threshold was determined as the minimum of the square root of the sum of squared ROC coordinates, resulting in a final threshold of 0.13.

**Figure 3.**
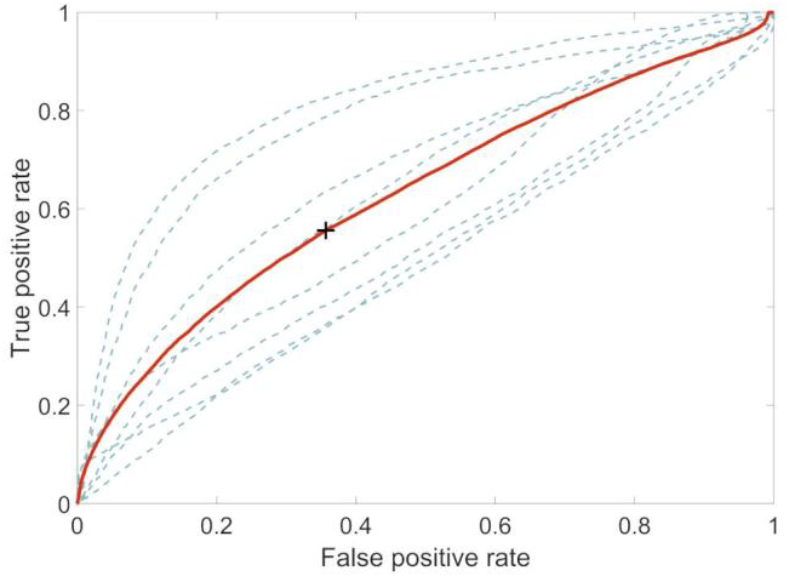
ROC curves per participant (blue dashed lines) show true positive activations in task-specific muscles (flexors for dorsal flexion task; extensors for plantar flexion task) and false positives in other muscles. A final aggregated ROC curve (red line) determines the optimal ReHo threshold (black cross) across all subjects.

Statistically significant differences were observed in the tibialis anterior (TA), extensor digitorum longus (EDL), and soleus muscles (SO). During plantar flexion compared to dorsal flexion, ReHo values in TA and EDL were lower. In contrast, the percentage values in the soleus increased during plantar flexion relative to dorsal flexion. The results of the statistical groupwise comparison are shown in Figure 4.

**Figure 4.**
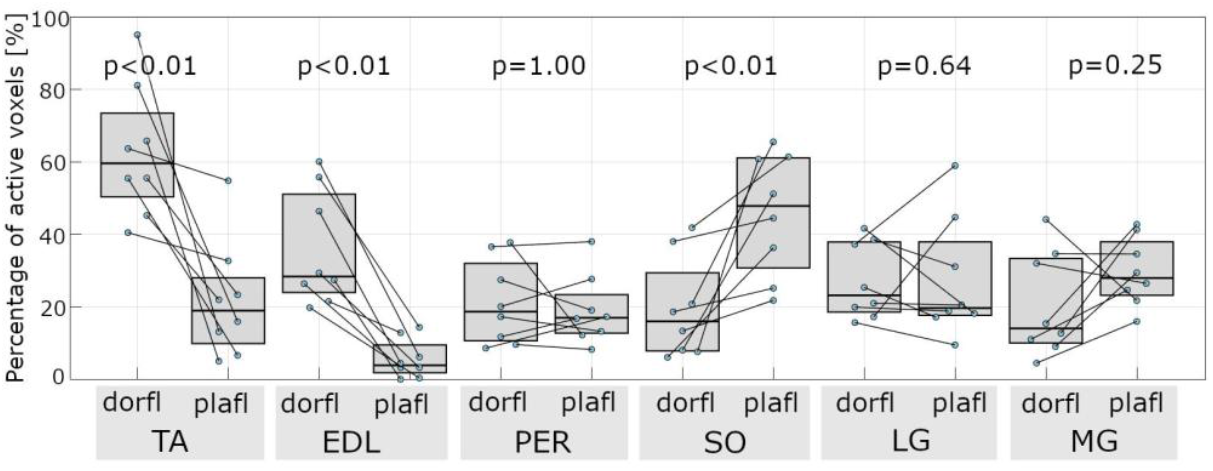
Percentage of active voxels (based on zReHo) in each segmented muscle between dorsal flexion (dorfl) and plantar flexion (plafl). A pairwise Wilcoxon signed-rank test was used for the statistical analysis. Blue points represent the percentage increase in zReHo values for each subject. Values corresponding to the same subject are connected by a black line. TA – tibialis anterior, EDL – extensor digitorum longus, PER – peroneus longus, SO – soleus, LG – lateral gastrocnemius, MG – medial gastrocnemius;A-anterior, P-posterior, L-left, R-right.

Through a visual comparison of resting and activated muscle behavior, differences in zReHo parameters were observed in all subjects (See Figure 5 with the main calf area of each subject). In individual analysis, it was observed that during plantar flexion, zReHo exhibited the highest values in the soleus muscle. Meanwhile, the gastrocnemius muscles (both medial and lateral) showed activity in only a subset of participants rather than uniformly across the group. On occasion, there was also activity observed in the flexor hallucis longus region, though this particular muscle was not specifically segmented and assessed in this study.

**Figure 5.**
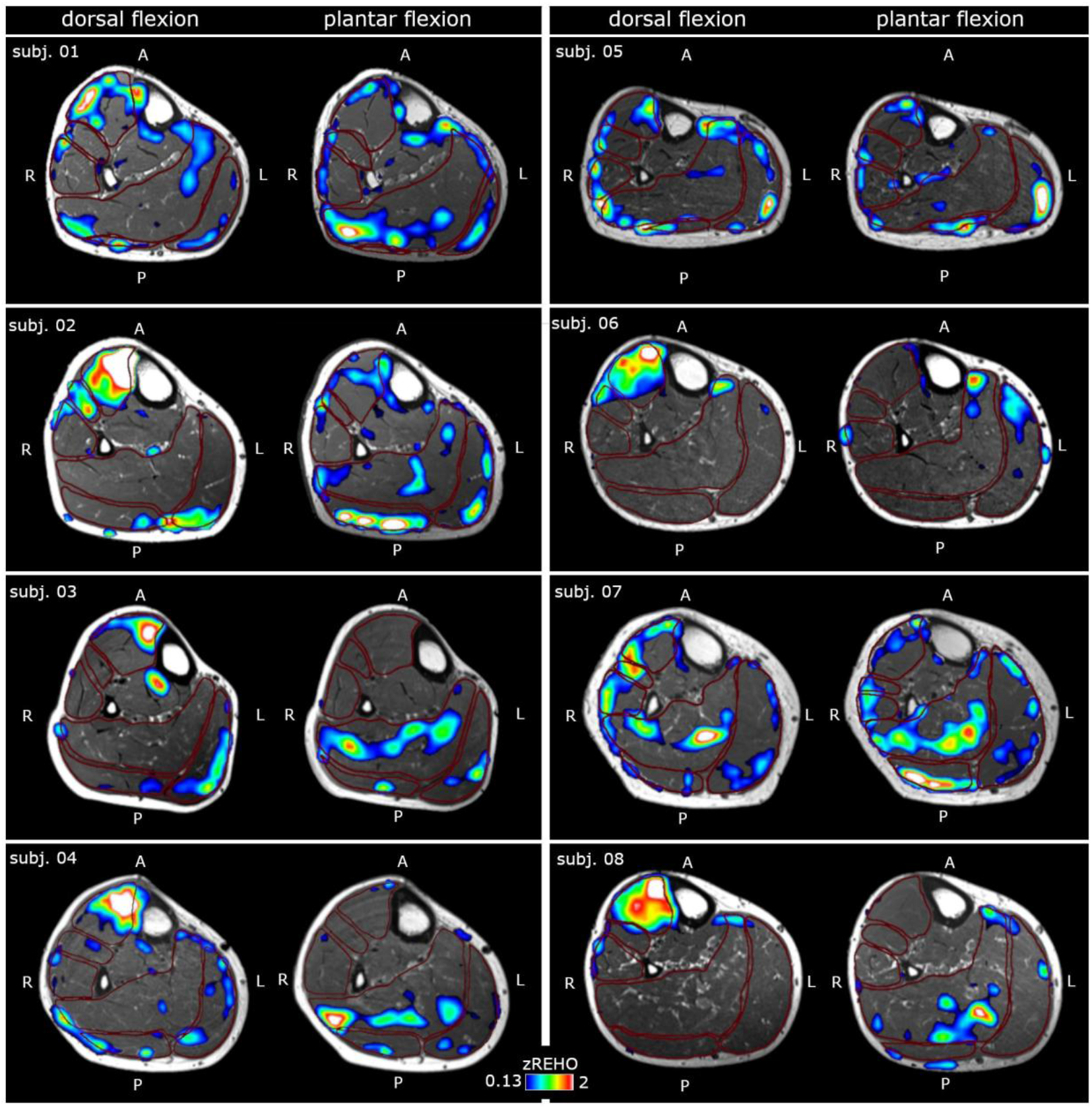
zReHo results in calf muscles during dorsal flexion (left) and plantar flexion (right) for each proband. The slices were obtained at the largest circumference of the anterior compartment. The zReHo scale (0.13-2) was the same for all probands and is shown bellow the slices. The zReHo threshold was determined using ROC analysis (see Methods for details). Probands are organized in two columns. Anatomical orientation is indicated as follows: A-anterior, P-posterior, L-left, R-right.

During dorsal flexion, an increase in ReHo was observed in the tibialis anterior muscle across all participants. Additionally, two participants showed an increase in ReHo parameter in the soleus and the medial gastrocnemius muscle during dorsal flexion.

## Discussion

Significant differences in ReHo values were found within the same muscle during plantar and dorsal flexion. The analysis of zReHo active voxel percentages revealed notable changes in the tibialis anterior, extensor digitorum longus, and soleus muscles. These differences were not only quantifiable but also visually distinguishable, further supporting the validity of resting-state fMRI (rs-fMRI) as a tool for mapping muscle activity.

An increase in zReHo values was observed in the active muscles compared to the inactive ones. However, this increase was not uniformly distributed across the entire muscle, aligning with previous research indicating intramuscular inhomogeneity (Akima et al., 2000). These findings suggest that different regions of the same muscle may engage to varying extents depending on specific biomechanical demands.

To capture this variability more accurately, we segmented the entire muscle belly, following approaches used in several studies (Mendiguchia et al., 2013; Zaeske et al., 2022). This allowed detection of all activity within the muscle, particularly when activation patterns are inconsistent across regions. In contrast, using only a spherical region of interest, as done in some previous studies (Bourne et al., 2017; Bourne et al., 2018), may miss relevant localized activations and underestimate true muscle engagement.

During plantar flexion, the highest zReHo values were consistently observed in the soleus muscle, indicating robust and uniform activation of this muscle across participants. Occasional activity was also noted in the region of the flexor hallucis longus; however, this muscle was not specifically segmented or evaluated in the present study. In contrast, the medial and lateral gastrocnemius muscles exhibited increased ReHo in only a subset of individuals, indicating a more variable and possibly participant-dependent contribution to plantar flexion.

During dorsal flexion, elevated zReHo was consistently detected in the tibialis anterior muscle across all participants, reinforcing its primary role in this movement. Interestingly, several individuals also showed increased ReHo in the soleus and medial gastrocnemius muscles. This activity may have been induced by passive muscle stretching. Notably, the stretching of these muscles during dorsal flexion is not a purely passive process and may elicit neuromuscular responses that resemble activation. However, these responses were comparatively less pronounced and more spatially limited than those observed in actively contracting muscles.

Elevated ReHo values in active muscle regions likely reflect increased local synchronization of BOLD signal fluctuations driven by coordinated motor unit recruitment and enhanced perfusion. During muscle contraction, spatially clustered motor activity and consistent neurovascular responses produce temporally aligned signal changes, resulting in higher regional homogeneity. This interpretation aligns with findings from brain studies, where ReHo increases in functionally engaged areas (Zang 2004).

Compared to brain imaging, fMRI analysis of muscle presents unique challenges, primarily due to variability in muscle shape caused by body position. The brain remains stable within the skull, whereas muscle morphology may shift. To address this variability, a single anatomical volume is acquired alongside the functional set, capturing muscle shape in the specific scanning posture. This pilot study highlights the potential of rs-fMRI to identify contracted muscle groups without relying on activation paradigms. By assessing spontaneous BOLD signal fluctuations, rs-fMRI enables the evaluation of muscle activity using only one position and one set of functional volumes. This approach simplifies the imaging process and enhances time efficiency, making it a practical tool for both clinical and research applications.

This study has several limitations. First, the sample size was small, and the results should be interpreted as preliminary. The results found must be supported and validated within the study with a larger number of probands. Second, we did not use electromyography concurrently with MRI to validate muscle activity. Instead, muscle activation was inferred based on anatomical knowledge. Nevertheless, within this limited sample, our rs-fMRI protocol successfully distinguished active muscle groups from inactive ones in individual analysis and offers new possibilities for assessing active muscle groups without the use of activation paradigms utilizing BOLD rs-fMRI.

To our knowledge, no previous studies have systematically attempted to use rs-fMRI for evaluating sustained muscle contraction patterns. One possible reason for this gap is the longstanding assumption that muscle BOLD signals are more susceptible to artifacts and less reliable than cerebral BOLD. Our findings challenge this view and suggest that reliable functional information can be extracted even in skeletal muscle.

If replicated and refined, this method could be applied clinically in cases where task-based paradigms are not feasible, such as in patients with involuntary or pathological muscle activation. An example of such a disease is cervical dystonia characterized by sustained, involuntary neck muscle contractions. Accurately identifying these hyperfunctioning muscles is essential for effective treatment, such as targeted botulinum toxin injections. Rs-fMRI may enable objective mapping of overactive muscle groups, including deep muscles that are difficult to assess non-invasively, thus serving as a valuable complement to electromyography.

In summary, our findings confirm the feasibility of detecting muscle activity from a single resting-state fMRI session and highlight the potential of this neuroimaging technique for novel diagnostic and therapeutic applications in neuromuscular disorders, especially when conventional paradigms cannot be employed.

## Data Availability

All data produced in the present study are available upon reasonable request to the authors.

## Acknowledgment

This study was supported by the National Institute for Neurological Research (Programme EXCELES, ID project no. LX22NPO5107) - Funded by the European Union - Next Generation EU.

Special thanks go to Květoslav Vtípil and Radovan řáček for their assistance with MRI data acquisition at Na Homolce Hospital.

## Notes

### Competing Interest Statement

The authors have declared no competing interest.

### Author Declarations

Ethics committee of General University Hospital in Prague gave ethical approval for this work.

